# Geospatial distribution of Hepatitis B prevention services in Wakiso District, Central Uganda

**DOI:** 10.1101/2022.04.20.22274066

**Authors:** Tonny Ssekamatte, John Bosco Isunju, Aisha Nalugya, Solomon Tsebeni Wafula, Rebecca Nuwematsiko, Nakalembe Doreen, Winnifred K. Kansiime, Muyanga Naume, Joana Nakiggala, Justine N. Bukenya, Richard K. Mugambe

## Abstract

**Introduction:** Despite global and national efforts in place for the prevention and control of Hepatitis B, there remains a gap in access to hepatitis B prevention services such as testing and vaccination. Nonetheless, there is limited evidence of the geospatial distribution of Hepatitis B services. This study established the geospatial distribution of HBV vaccination services in Wakiso District, Uganda.

**Materials and methods:** A cross-sectional quantitative descriptive study was conducted among 55 healthcare facilities including 6 hospitals, and 49 primary care facilities in Wakiso district. Data were collected using the KoboCollect application. Quantitative data were analysed using STATA 14.0. A chi-square test was performed to establish the relationship between healthcare facility characteristics and the availability of hepatitis B services. ArcGIS (version 10.1) was used for analysis of geospatial data.

**Results:** The hepatitis B vaccine was available in only 27.3 % (15) of the facilities, and 60% (33) had testing services. Receipt of the hepatitis B vaccine doses in the last 12 months was associated with the level of healthcare facility (p=≤0.001) and location (p=0.030). Availability of the Hepatitis B vaccines at the time of the survey was associated with the level of healthcare facility (p=0.002) and location (p=0.010). Availability of hepatitis B testing services was associated with level of healthcare facility (p=0.031), ownership (p≤0.001) and location (p=0.010). Healthcare facilities offering vaccination and testing services were mostly in urban healthcare facilities, and close to Kampala, Uganda’s capital.

**Conclusion:** Hepatitis B services were sub-optimal across all healthcare facility levels, locations, and ownership. The majority of the hepatitis B prevention services were provided in urban settings, close to major towns, municipalities, and the city. This calls a extension of hepatitis B prevention services to rural, public and PNFP healthcare facilities.

## Introduction

Hepatitis B virus (HBV) infection remains a serious global public health concern, affecting over 257 million people (1, 2). Low-and-middle income countries (LMICs) bare the greatest burden of HBV infection, accounting for almost all (96%) individuals living with the disease (1). The incidence of HBV infection is 9.2 times higher in LMICs than high-income countries (1). The WHO African region has the second highest prevalence of HBV infection (6.1%) while America has the lowest (0.7%) (1). The prevalence of HBV infection in Uganda is 4.1% among adults and 0.6% among children. It is highest in the mid-north (4.6%) and lowest in the south-western region (0.8%) of the country (3). Despite the high public health burden of HBV infection in LMICs including Uganda, access to testing, vaccination, treatment and management services remains suboptimal (1). Nonetheless, a large burden of chronic infections requires increased access to testing and treatment services (4, 5).

HBV is an infection of the liver, transmitted through contact with blood or other body fluids of an infected person. The risk of chronic HBV infection and co-infection is high among under-fives, health care providers, and people living with HIV/AIDS respectively (1, 6). A large proportion of HBV cases are carriers who may remain asymptomatic for a long time, thus increasing their risk of liver cirrhosis, hepatocellular carcinoma and early death (1). Access to vaccination and screening/testing is critical in the prevention and management, and elimination of HBV infection by 2030 (1, 7). Availability of testing services is key in the diagnosis of chronic HBV, and consequently linkage of patients to appropriate care and treatment, which eventually delays progression of liver disease (4, 5, 7). Additionally, availability of testing services provides an avenue for counselling on risky behaviours, provision of prevention commodities such as sterile needles and syringes, and vaccination (4, 5). Vaccination reduces the risk of HBV infection, and co-infection with Hepatitis D virus, which is known to aggreveate the outcome of HBV infection (1, 8, 9).

Although evidence on global and regional coverage of hepatitis B vaccination among adults is scarce, high-income countries with vaccination data reveal low rates (10). Limited access to testing and vaccination services among adults born before the HBV vaccination era increases their risk of infection and death (1, 5). Available data on vaccination coverage among groups at an elevated risk of the HBV infection in low-income countries reveal suboptimal rates (11-14).

There have been various strategies at global, regional and national level aimed at reducing disparities in access to hepatitis B prevention, care and treatment services (15). In 2016, the World Health Organisation (WHO) developed a global health sector strategy on viral hepatitis (2016–2021) which envisioned a world where viral hepatitis transmission is halted and everyone living with viral hepatitis has access to safe, affordable and effective prevention, care and treatment services (1, 7). In response to the WHO recommendations and targets, the Ugandan Ministry of Health (MoH) called for increment in massive community awareness of HBV, screening and vaccination of all susceptible persons, improving the supply chain for HBV related commodities and supplies, attachment of patients to treatment care, and support programs, integration of services and resource mobilisation and equipping Healthcare practitioners (HCPs) with adequate knowledge on the management of HBV infection (MoH, 2016b). More so, the government of Uganda allocated US$2,847,000 in 2016 towards the procurement of vaccines, laboratory reagents, and anti-viral drugs for the prevention and treatment of Hepatitis B (16).

Hepatitis B vaccination in Uganda was introduced in 2002, implying that adults born before that year missed out on the routine vaccination given to newborns at 6, 10 and 14 weeks of age through the expanded programme on immunisation (17). Despite efforts to improve access to hepatitis B prevention, care and treatment services, their distribution in Wakiso district is unknown. (17). Our study sought to undertake a geospatial and descriptive analysis of hepatitis B prevention services, vaccine doses, testing kits and trained personnel, as well as understand the relationship between healthcare facility (HCF) characteristics and availability of hepatitis B prevention services and supplies.

## Materials and methods

### Study setting

This study was conducted in Wakiso district, Central Uganda in July 2018. Wakiso District is located in the Central Region of Uganda and partly encircles Kampala, Uganda’s capital. Wakiso District boarders Mpigi, Luweero, Nakaseke and Mityana Districts in the North; Mukono in the East and Kalangala district to the South. It has a total of 7 health sub-districts namely; Busiro east, Busiro north, Busiro south, Kyadondo east, Kyadondo north, Kyadondo south, and Entebbe municipality. In 2014, Wakiso was estimated to have a population of 1,997,418 people, of which 1,048,383 were females (18). The district has a total of 589 HCFs, among which 72 are owned and managed by the government (public), 477 are privately owned (private for-profit) and 40 are private not-for-profit (19). Based on the level of the HCFs, 234 are private clinics, 153 are health centre IIs, 165 are health centre IIIs, 19 are health centre IVs, 14 are hospitals, and 3 are specialised clinics. Wakiso is also home to the Entebbe regional referral hospital (19).

In terms of management of health services in Uganda, healthcare services in the district are overseen by the District Health Officer who is supported by two assistants. The district is further divided into health sub-districts, which are overseen by a management committee. The functions of the committee include monitoring the general administration of the health Sub-district on behalf of the district local council and MoH. Each of these committees has a health sub-district manager, often an in-charge at a health centre IV, whose role involves coordination, planning, and budgeting for medical supplies and infrastructure, budget execution, and supervision of lower-level HCFs such as health centre IIIs and IIs (20).

### Study design and data collection tools

A cross-sectional study design was used to collect quantitative data from each of the study HCFs. A structured questionnaire was used to obtain data from the HCF managers. Where observations were required, questions prompting the researcher to make such observations were embedded in the data collection tool. The structured questionnaire was developed by the lead investigators (TS, JBI, and RKM) after a thorough review of literature pertaining provision of vaccination services (21, 22), and the service availability and readiness assessment (SARA) framework. The SARA framework is designed as a systematic survey to generate a set of core indicators of services, which can be used to measure progress in health system strengthening over time (23, 24). The study specifically focused on service availability, which is defined as the physical presence of the delivery of services, encompassing the health infrastructure, core health personnel, and service utilization (23, 24). The SARA is useful in the identification of the proportion of HCFs offering a particular intervention/service-in this case hepatitis B vaccination services, and whether the HCFs offering the service (HBV services) have the minimum standard in terms of equipment, trained staff and guidelines, diagnostic capacity, and medicines to provide an adequate level of service (service readiness) (23, 25). The data collection tool was reviewed and evaluated for face and internal validity by experts in viral hepatitis research, based at the Makerere University College of Health Sciences.

### Study variables

The validated tool captured data on the HCF characteristics such as level (categorised as hospital, health centre IVs, and IIIs), ownership (Private for-profit (PFP), Private not-for-profit (PNFP), and Public/government-owned), and location (rural/urban). Urban HCFs included those administratively located in municipalities or town councils while rural HCFs were those located in a sub-county (26). Data were collected on the availability of vaccination and testing services, trained personnel, and vaccination storage equipment such as refrigerators. In addition, observations were made to ascertain the availability of hepatitis B testing kits, vaccine doses, infection prevention and control promotion materials, and disposal of medical wastes such as sharps.

### Sample size, sampling procedure, data collection procedures, and data analysis

A total of 55 HCFs were surveyed. The sampling of the HCFs in the current study was purposive and has been detailed in our earlier publications (11, 27). Briefly, based on the national health facility inventory (19), we purposively selected 6 hospitals, 16 health centre IVs and 33 IIIs because these provided high-risk medical interventions such as blood transfusions, delivering higher numbers of mothers, and other surgical procedures that can elevate the risk of transmission of HBV (27, 28). Upon selection of the HCFs, data were collected using the Kobo Collect mobile application which was installed on Android-enabled mobile phones and tablets, and preloaded with the electronic version of the questionnaire. Research Assistants were required to upload the data onto a cloud server daily to avoid any risk of losing it. Upon completion of the survey, data were downloaded in the MS EXCEL format, checked for consistency, and cleaned. Afterwards, data were exported to STATA version 14.0 for analysis. Cross tabulations based on the level of the HCF, ownership, and location were done and are presented in the current paper. The Chi square (χ^2^) test was performed to establish the relationship between HCF characteristics and availability of hepatitis B related services and supplies.

### Geospatial distribution of prevention services

The digital data collection tool which was used to gather data from HCFs also picked Geographical Positioning Systems (GPS) coordinates, i.e., northings and eastings. Location data were stored on the cloud server hosted by Kobo collect, together with attribute data about the HCFs. The data set was later downloaded as an excel file, checked for inconsistencies, and cleared of any errors. The data file was then saved as a comma delimited text file and imported as a layer into a GIS environment using ArcGIS (version 10.1). From the imported layer, a shapefile of point plots and corresponding attribute tables were generated. The point plots were overlaid onto administrative boundaries of Wakiso district and its environs, obtained from the Ugandan Bureau of Statistics (UBOS). Subsequently, shapefiles for different variables were generated depending on the research questions that needed explicit representation. Maps showing the geospatial distribution of different attributes such as availability of HBV, and routine vaccination schedule for health workers were generated and exported as a jpeg.

### Quality control and quality assurance measures

Data were collected by 6 Research Assistants who were supervised by one of the lead investigators (TS). Before data collection, Research Assistants underwent a four days’ training to familiarise themselves with the data collection protocol. As part of the training, the data collection tools were pretested at two primary HCFs in peri-urban areas of Kampala City. The pre-test HCFs had comparable characteristics as those in the study district. These characteristics included conducting high-risk medical interventions such as conducting high volume deliveries, which increases occupational exposure to viral hepatitis.

### Ethics statement

The study was approved by the Makerere University School of Public Health Higher Degrees Research and Ethics Committee. Administrative clearance was sought from the Wakiso district Local government and the management of the participating HCFs. Written informed consent was obtained from the study respondents before interviews or observations could happen. All informed consent discussions were done in English since all health workers were conversant with the language.

## Results

### Background characteristics of health facilities

Out of the 55 facilities visited, most 61.8% (34) were health centres IIIs and 47.3% (26) were privately owned. Most of the HCFs visited were located in the urban setting 65.4% (36/55) and Busiro North sub-district had the highest number 29.1% (16/55) of the facilities visited (Table 1).

**Table 1:** Background characteristics of study healthcare facilities.

| **Variables** | **Category** | **Frequency (N=55)** | **Percentage (%)** |
| --- | --- | --- | --- |
| Level of health facility | Health Centre III | 34 | 61.8 |
|  | Health Centre IV | 17 | 30.9 |
|  | Hospital | 04 | 7.3 |
| Health facility Ownership | Public | 23 | 41.8 |
|  | Private | 26 | 47.3 |
|  | Private-not-for-profit | 06 | 10.9 |
| Location of the facility | Urban | 36 | 65.4 |
|  | Rural | 19 | 34.6 |
| Name of the Health sub-district | Busiro North | 16 | 29.1 |
|  | Busiro East | 7 | 12.7 |
|  | Busiro south | 5 | 9.1 |
|  | Kyadondo North | 10 | 18.2 |
|  | Kyadondo south | 06 | 10.9 |
|  | Kyadondo East | 10 | 18.2 |
|  | Entebbe municipality | 01 | 1.8 |

### Availability and distribution of Hepatitis B vaccine doses, testing kits and trained personnel

Only 29% (16) of the 55 HCFs reported having received hepatitis B vaccine doses in the last 12 months. At the time of the study, the hepatitis B vaccine was available in only 27.3 % (15/55) of the facilities; mostly in HC IVs 52.9% (9/17) and lowest in HC IIIs 11.8% (4/34). More urban HCFs, 93.3% (14/15) had hepatitis B vaccine available at the time of the study than rural ones 6.7% (1/14). Most HCFs 60.0% (33/55) had testing kits among which all hospitals and PNFP facilities had hepatitis B testing kits at their facilities. More urban HCFs, 72.2% (26/36) had testing kits than rural ones 36.8% (7/19). Although 30.9% (17/55) of the HCFs had personnel trained on Hepatitis B management; only 1 hospital 25.0% (1/4) and 3 13.0% (3/23) public facilities had their personnel trained on hepatitis B management. Infection control promotion materials were found in 89.1% (49/55) of the facilities; 91.2% (31/34) in HC IIIs, 82.4% (14/17) in HCIV and all hospitals. Nearly three quarters 65.6% (36/55) of the HCFs had post-exposure prophylaxis (PEP) guidelines in case of accidental injuries; this was lowest in public facilities 65.2% (15/23). Almost all facilities 96.4% (53/55) had handwashing facilities (Table2).

### Availability and distribution of routine vaccination services for healthcare providers

Only 29.1% (16/55) of the HCFs surveyed had a routine vaccination schedule for the health care providers. The majority of the HCFs that had a routine vaccination schedule were clustered in the same area, with just a few dispersed outwards (Figure 1). The majority of the HCFs were also located just adjacent to Kampala, Uganda’s Capital with more than half, 62.5% (10/16) in the urban areas and over a third, 37.5% (6/16) in the rural areas.

**Figure 1:**
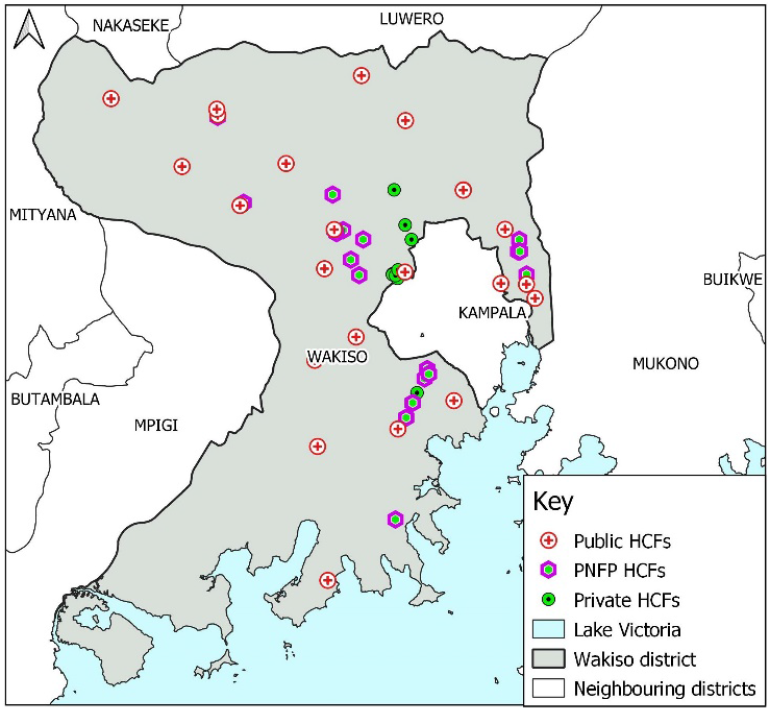
Map of Wakiso district showing the distribution of sampled healthcare facilities by ownership status.

**Figure 2:**
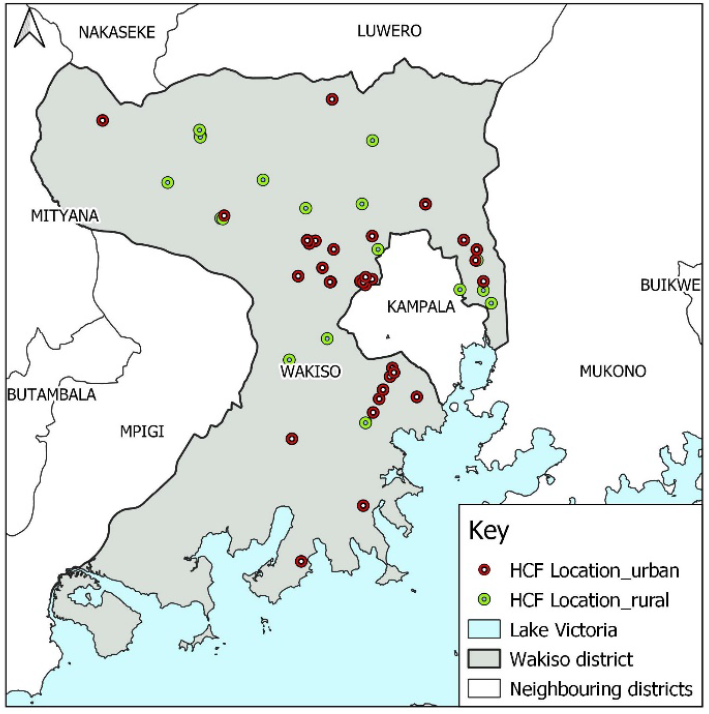
Map of Wakiso district showing the distribution of sampled healthcare facilities by location.

**Figure 3:**
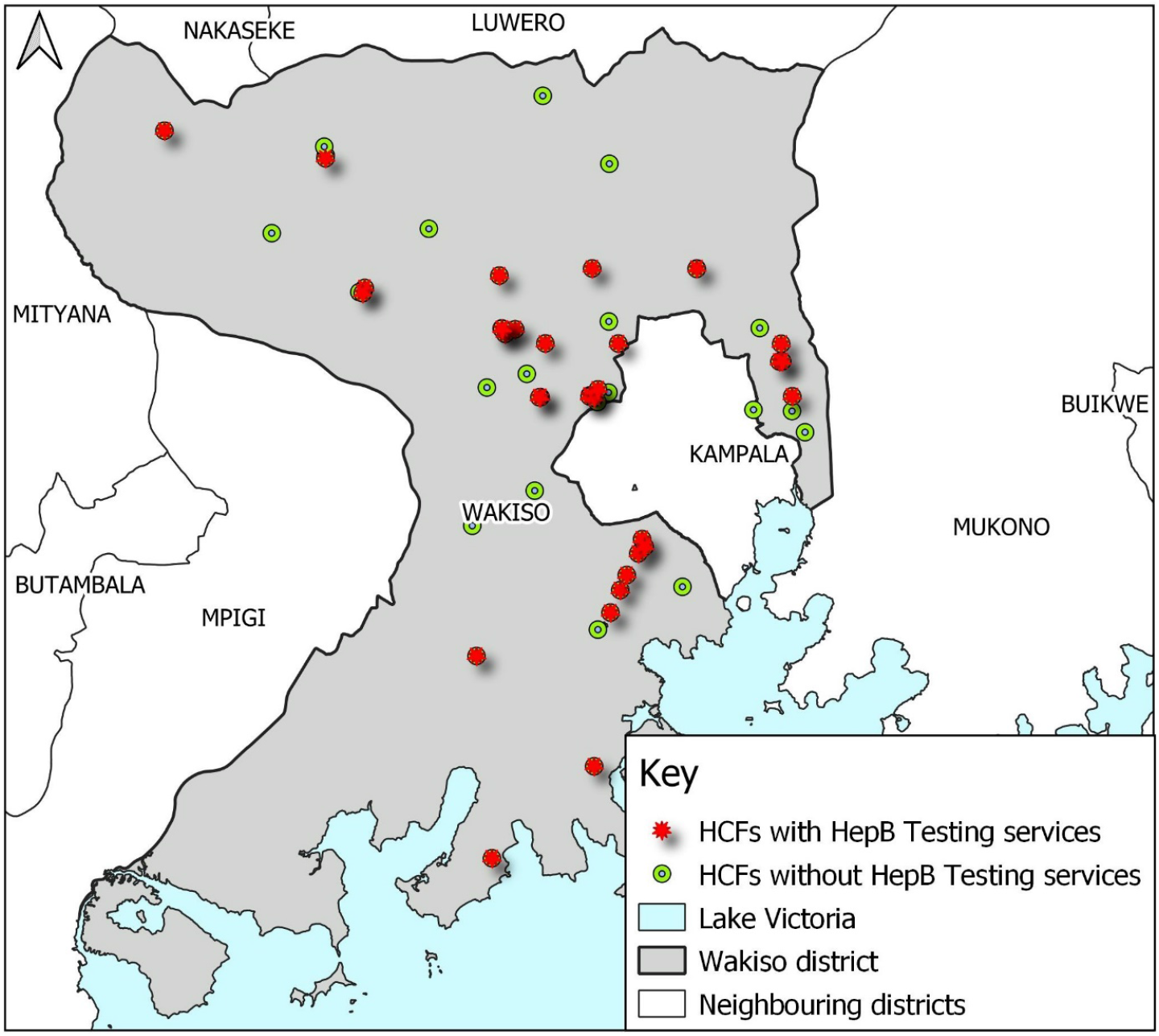
A map of Wakiso district showing the distribution of healthcare facilities that offer hepatitis B testing services.

**Figure 4:**
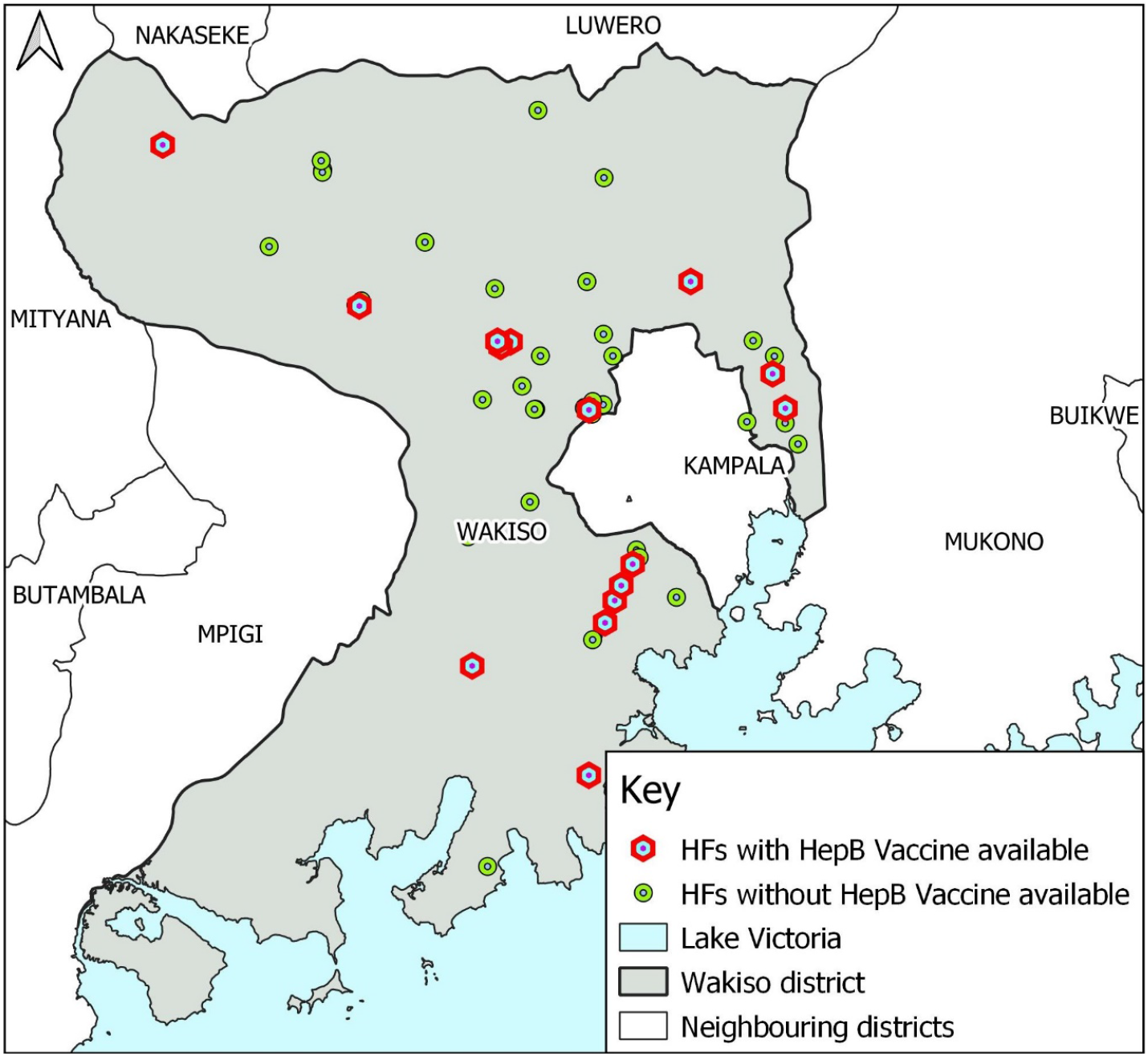
A map showing healthcare facilities with the hepatitis B vaccine in Wakiso district, Uganda.

**Figure 5:**
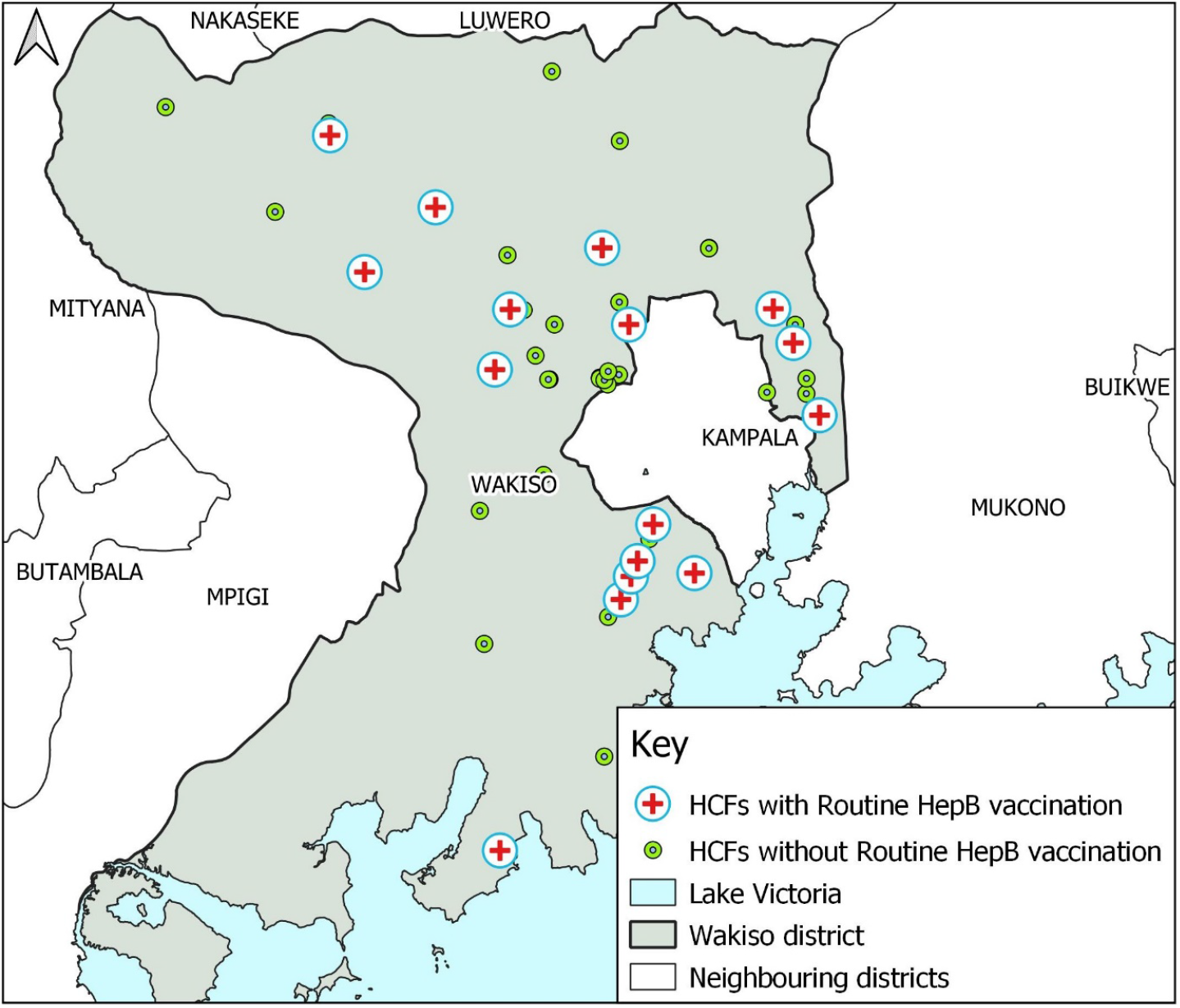
A map of Wakiso district showing healthcare facilities with a routine vaccination schedule (as either statatic or outreach) for healthcare providers and other high-risk groups.

### Relationship between healthcare facility characteristics and availability of hepatitis B prevention services and supplies

There was a statistically significant association between level of HCF and receiving hepatitis B vaccine doses in the last 12 months (p=≤0.001), having Hepatitis B vaccines at the time of the survey (p=0.002), availability of hepatitis B testing services (p=0.031), having hepatitis B testing kits in stock (p=0.013), having reminders and/or job aids that promote the reduction and use of injections, safe administration of injections, or safe disposal of used injection equipment available at this facility (p=0.003) and having color-coded waste bins to promote safe waste management (p=0.043) (Table 2).

**Table 2:**
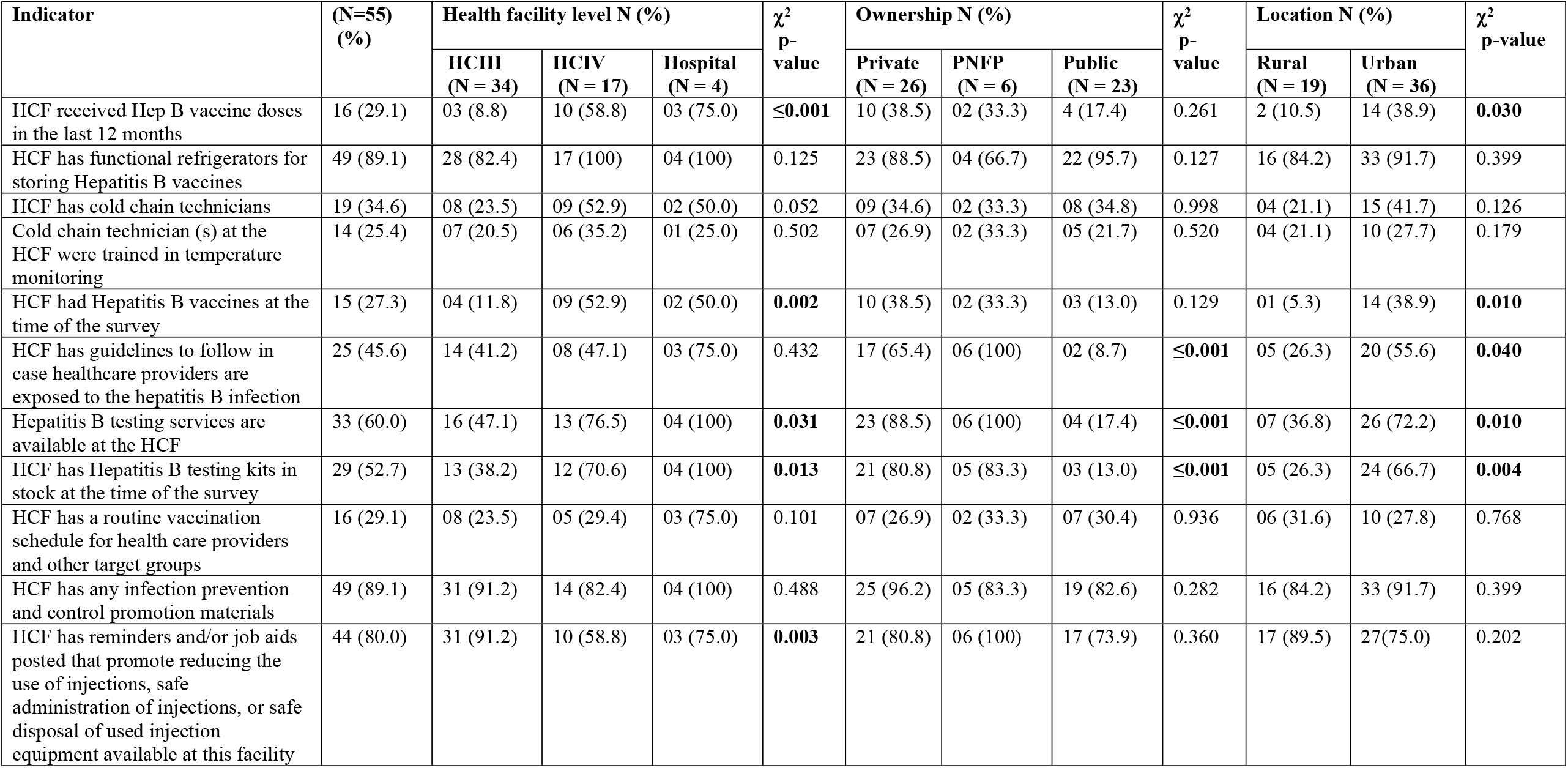
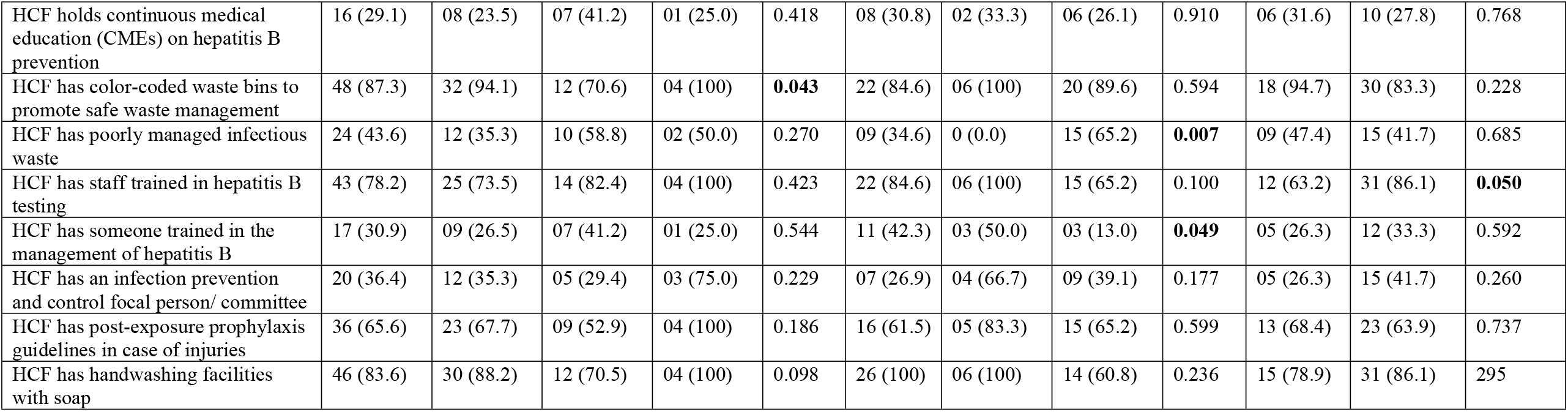
Availability of hepatitis B prevention services in Wakiso district, Central Uganda, stratified by level of the healthcare facility, ownership, and location.

There was a statistically significant association between ownership of the HCF and availability of guidelines to follow in case healthcare providers are exposed to the hepatitis B infection (p≤0.001), availability of hepatitis B testing services (p≤0.001), availability of hepatitis B testing kits (p≤0.001), poor management of infectious waste (p=0.007), and having someone trained in the management of hepatitis B (p=0.049).

The location of HCF was associated with receiving hepatitis B vaccine doses in the last 12 months (p=0.030), and availability of Hepatitis B vaccines (p=0.010), testing services (p=0.010) and Hepatitis B testing kits (p=0.004). The location of the HCF was also associated with availability of guidelines to follow in case healthcare providers are exposed to the hepatitis B infection (p=0.040) and presence of staff trained in hepatitis B testing (p=0.050).

## Discussion

The main aim of this study was to assess the availability and geospatial distribution of Hepatitis B prevention services in Wakiso District, Central Uganda. This is important since is unmasks variation in service availability across a geographic space. Our analysis found that only 29% of the HCFs had hepatisis B vaccine supplies in the last 12 months, but the vaccine was available in only 27.3 % of the HCFs at the time of the survey. We established that less than half of the HCFs had infection prevention and control (IPC) guidelines to follow when exposed to HBV infections, and only two-thirds of the HCFs had PEP guidelines in case of injuries. Geospatial analysis indicated that the majority of the HCFs offering routine HBV vaccination were clustered in the same area, with just a few dispersed outwards. Hepatitis B vaccination and testing services were mainly offered by health centre IVs and hospitals.

Analysis of the geospatial distribution of hepatitis B prevention services such as vaccination, screening, and testing, revealed that HCFs offering such services conformed to a nucleated pattern. HCFs were located close to each other. Many of these HCFs were located close to Kampala, Uganda’s capital, and in municipalities and major town councils. The location or proximity of HCFs providing Hepatitis B prevention services to the more urbanised areas is not surprising. Urbanised areas are more likely to have a higher demand for services compared to the less urbanised or rural settings. There is also evidence that urban dwellers are more likely to be knowledgeable about their health, and thus can demand services. However, this has implications for hepatitis B prevention. It is known that rural dwellers in Uganda have lower incomes compared to urban dwellers (29, 30). Therefore, the unequal distribution of hepatitis B prevention services is likely to limit their uptake.

Our study revealed that only 29% of the HCFs reported having hepatitis B vaccine doses in stock in the last 12 months. Our findings indicate that less than a third of the HCFs had hepatitis B vaccines in stock during the study period (2019). The low proportion of HCFs with hepatitis B vaccine doses in stock during the study period may have been attributed to the low prevalence of the disease in the region. The central region, where Wakiso district is located had a lower prevalence (1.6%) of hepatitis B compared to other regions (4.6% in mid-north, 3.8% in West Nile, 4.4% in northeast among other regions) (31, 32). Due to the low prevalence of hepatitis B in the central region, Wakiso district was in 2015 not prioritised by the Ugandan Ministry of Health for mass hepatitis B prevention services including screening, vaccination of adults, and testing (32). The low prevalence and consequently low prioritisation of hepatitis B prevention services by the Ugandan Ministry of Health may have driven the availability and distribution of hepatitis B services in HCFs in Wakiso district. Besides, the low demand for hepatitis B prevention services, which resulted from limited knowledge and a negative attitude towards hepatitis B prevention among healthcare providers in Wakiso district, as earlier reported by Ssekamatte, Isunju (27) could also explain the limited coverage in HCFs. The study reported that 42.2% of healthcare providers exhibited poor knowledge of HBV infection transmission and prevention, 41.8% had a negative attitude while 41.5% exhibited poor hepatitis B prevention practices. There is evidence that unfavourable knowledge, attitude, and practices can affect the demand and supply of healthcare services (33).

A higher proportion of PNFP and private HCFs had hepatitis B prevention services such as screening and testing compared to the public HCFs. This is not surprising given the fact that procurement of the vaccine greatly depended on the demand, and that the vaccine was paid for by the clients. Therefore, the provision of hepatitis B vaccination services such as testing and vaccination was seen as a business from which PNFP and private HCFs would make profits. There is evidence of HCFs providing services only if they can make a profit (34). The low proportion of public HCFs reporting having hepatitis B vaccines in stock in the last 12 months could be attributed to the fact that Wakiso district was not in a high-burden region, and thus, not prioritised. The Ugandan Ministry of Health rolled out free vaccinations in the country starting with regions of high prevalence that did not include Wakiso district. Thus public/government HCFs in low burden areas did not have the vaccine readily available (32).

Less than half of the HCFs in our study setting had guidelines patients and healthcare providers could follow when exposed to the hepatitis B virus. In addition, about two-thirds of the HCFs had PEP guidelines in case of injuries. The low proportion of HCFs with infection prevention and control guidelines could have been due to the limited investment in hepatitis B prevention and control to support development and distributribution of guidelines, policies and standards to HCFs. Limited funding is widely reported to impact the provision of healthcare services, and the development and implementation of guidelines and policies (35).

Less than a third of the study HCF held continuing medical education sessions(CMEs) on hepatitis B prevention. The CMEs helps healthcare providers to maintain, develop, or increase the knowledge and skills about the provision of healthcare. This is a considerably low proportion given the public health importance of hepatitis B. The low proportion of HCFs holding CMEs may have resulted from the fact that less than a third of the HCF had a healthcare provider trained in the management of hepatitis B, despite more than three quarters being trained in hepatitis B testing. The fact that only a few HCFs had healthcare providers trained in hepatitis B management indicates a knowledge gap among healthcare providers in HCFs in Wakiso district. Therefore, our study recommends the need to train healthcare providers, particularly those in health centre IIIs and public HCFs, irrespective of the location of HCF.

## Conclusions

Our study revealed an uneven distribution of hepatitis B services. The majority of the hepatitis B services were clustered around major towns, municipalities, and Kampala City, with just a few sparsely located in rural areas. Regarding the availability of hepatitis B prevention services, less than a third of the HCFs had hepatitis B vaccine doses in stock in the last 12 months, less than a third had a routine vaccination schedule for the health care providers while sightly more than half offered hepatitis B testing services. The proportion of HCFs offering hepatitis B services was higher among urban HCFs, private HCFs, and hospitals. This calls a extension of hepatitis B prevention services to rural, public and PNFP healthcare facilities.

## Data Availability

All relevant data are within the paper and its supporting information files.

## Declarations

Consent for publication: Not Applicable

## Availability of data and material

All relevant data are within the paper and its Supporting Information files.

## Competing interests

The authors declare that they have no competing interests.

## Funding

No funding source to declare.

## Authors’ contributions

TS, JBI and RKM were involved in the conceptualisation of the study, design, data analysis, and drafting of the manuscript. AN STW, RN, ND, MN, JN, WKK and JNB participated in data analysis, interpretation of findings and drafting of the manuscript. All authors read and approved the final manuscript. JBI drew the maps presented in the manuscript.

## Acknowledgements

We thank the Wakiso District Health office for granting the study team administrative clearance during the survey. Our sincere thanks also go to the management/administration of the different healthcare facilities, without whom, this study would not have been accomplished. Finally, we appreciate the research assistants for their role in undertaking the survey from which the current manuscript has been written.

## Notes

### Competing Interest Statement

The authors have declared no competing interest.

### Author Declarations

The study was approved by the Makerere University School of Public Health Higher Degrees Research and Ethics Committee.

